# Adverse events of special interest for COVID-19 vaccines - background incidences vary by sex, age and time period and are affected by the pandemic

**DOI:** 10.1101/2021.10.04.21263507

**Authors:** Fredrik Nyberg, Magnus Lindh, Lowie E.G.W. Vanfleteren, Niklas Hammar, Björn Wettermark, Johan Sundström, Ailiana Santosa, Brian K. Kirui, Magnus Gisslén

## Abstract

**Background:** With large-scale COVID-19 vaccination implemented world-wide, safety signals needing rapid evaluation will emerge. We report population-based, age- and-sex-specific background incidence rates of conditions representing potential vaccine adverse events of special interest (AESI) for the Swedish general population using register data.

**Methods:** We studied an age/sex-stratified random 10% sample of the Swedish population on 1 Jan 2020, followed for AESI outcomes during 1 year, as the COVID-19 pandemic emerged and developed, before the start of vaccinations. We selected and defined the following outcomes based on information from regulatory authorities, large-scale adverse events initiatives and previous studies: aseptic meningitis, febrile seizure, Kawasaki syndrome, MISC, post-infectious arthritis, arthritis, myocarditis, ARDS, myocardial infarction, stroke, ischemic stroke, hemorrhagic stroke, venous thromboembolism, pulmonary embolism, kidney failure, liver failure, erythema multiforme, disseminated intravascular coagulation, autoimmune thyroiditis, and appendicitis. We calculated incidence rates stratified by age, sex and time period (quarters of 2020), and classified them using Council of International Organizations of Medical Sciences (CIOMS) categories: very common, common, uncommon, rare, or very rare.

**Results:** We included 972,723 study subjects, representing the Swedish national population on 1 Jan 2020. We found that AESI incidence rates vary greatly by age and in some cases sex. Several common AESIs showed expected increase with age, while some (e.g. appendicitis, aseptic meningitis, autoimmune thyroiditis, Kawasaki syndrome and MISC) were more common in young people, and others exhibited a flatter age pattern (e.g. myocarditis, DIC and erythema multiforme). Consequently, the CIOMS classification for AESIs varied widely according to age. Considerable variability was suggested for some AESI rates across the 4 quarters of 2020, potentially related to pandemic waves, seasonal variation, healthcare system overload or other healthcare delivery effects.

**Conclusion:** Age, sex, and timing of rates are important to consider when background AESI rates are compared to corresponding rates observed with COVID-19 vaccines.

## Introduction

The novel severe acute respiratory syndrome coronavirus 2 (SARS-CoV-2) first emerged in Wuhan in December 2019 (1), causing the enigmatic disease subsequently named Coronavirus disease 2019 (COVID-19). During 2020, the disease developed into a large-scale pandemic, spreading to most countries in the world, including entering second, third or even fourth wave in many places, with cumulative numbers of over 83 million cases and 1.8 million deaths reported globally during the first year until end of 2020 (2). By the end of 2020, vaccines had been developed and vaccination campaigns were initiated in multiple countries, including Sweden. The first COVID-19 case in Sweden was reported on 31 January 2020, and in parallel with Europe becoming the epicenter for the epidemic during the Spring of 2020, the disease rapidly expanded in Sweden, causing severe strain on society and the healthcare system (3, 4). At the peak of the second big wave of infections, vaccination was initiated in the elderly population just before the end of 2020, and has continued at pace during 2021, reaching successively larger sections of the population and younger population groups.

With the initiation of vaccination against COVID-19 in Sweden and across the world by end of 2020, regulatory agencies, pharmaceutical companies, academia and other stakeholders are developing safety surveillance strategies, largely based on observational data. In Europe, for instance, the vACCine covid-19 monitoring readinESS (ACCESS) project funded by the European Medicines Agency includes a protocol to assess background rates of conditions considered to represent potential Adverse Events of Special Interest (AESI) from observational data (5). Similarly, The US FDA Center for Biologics Evaluation and Research have published a protocol on “Background Rates of Adverse Events of Special Interest for COVID-19 Vaccine Safety Monitoring” (6). Local background rates are generally considered most appropriate for obvious reasons (7), but expected rates can also be better informed by availability of background rates from many different populations, time periods and areas, to understand variability and consistency of rates. Data on background rates of AESI for COVID-19 vaccines from some different areas have started to appear in the scientific literature, for example a multi-database study by the Observational Health Data Sciences and Informatics (OHDSI) community (8). These efforts are important, as a comprehensive and thorough monitoring of vaccine safety will be an essential component in addressing public concerns about the rapid COVID-19 vaccine development and deployment, and support efforts to address vaccine hesitancy. Here, we provide descriptive epidemiology on background rates for a range of AESI in Sweden, looking at time periods just prior to and during the COVID-19 pandemic, and before extensive vaccination was initiated, in order to inform surveillance efforts in Sweden and internationally.

## Methods

### Study design

Population-based cohort study to estimate incidence rates of conditions representing Adverse Events of Special interest (AESI).

The analyses were based on the larger SCIFI-PEARL (Swedish Covid-19 Investigation for Future Insights – a Population Epidemiology Approach using Register Linkage) project, which has been described elsewhere (9). A subproject known as RECOVAC (Register-based large-scale national population study to monitor Covid-19 vaccination effectiveness and safety) focuses on COVID-19 vaccination research questions and this analysis is a first component of that effort.

### Data sources

The current analyses were based on the Swedish national population on 1 Jan 2020, before the outbreak of the COVID-19 pandemic. The study population was identified from national registers at Statistics Sweden, with age and sex data. To this study population we linked data from the National Patient Register (NPR), which includes data on all hospitalizations and outpatient specialist care visits in Sweden, including date of admission or date of visit and diagnoses coded with International Classification of Diseases version 10 (ICD-10) codes, Swedish national version (9).

### Study Population and study period

The study population was an age-sex stratified sample of the Swedish population as of 1 Jan 2020. The number of individuals sampled and sampling fractions from the total population (i.e. representing weights to obtain numbers representative for the general Swedish population) are given in Suppl Table 1. The population was followed for 1 year from the index date of 1 Jan 2020, i.e. during the quarter before the pandemic had spread substantially in Sweden and then during the first 3 quarters of pandemic spreading.

### Outcomes

The investigated outcomes represent adverse events of special interest (AESIs) that have been suggested as being particularly important to track following COVID-19 vaccination, and for which there is a need to have solid information on background rates. The AESIs were identified from the “Priority list of adverse events of special interest: COVID-19” AESI list by the Brighton Collaboration (10), the “Background Rates of Adverse Events of Special Interest for COVID-19 Vaccine Safety Monitoring” protocol published by the FDA Center for Biologics Evaluation and Research (6), and lists of AESI in protocols from the EMA ACCESS project (5). Algorithms defining AESIs by ICD-10 codes were developed based on these sources, some other study protocols and proposals, and a review of suitable codes in the Swedish national version of ICD-10. Events were identified by ICD codes from the NPR, representing hospitalization or outpatient specialist visit for the condition. Based on the availability of ICD codes in our database, the following conditions could be identified with all required ICD codes that we had defined: aseptic meningitis, febrile seizure, Kawasaki syndrome, MISC, postinfectious arthritis, arthritis (broad definition), myocarditis, ARDS, myocardial infarction, stroke, ischemic stroke, hemorrhagic stroke, venous thromboembolism, pulmonary embolism, kidney failure, liver failure, erythema multiforme, disseminated intravascular coagulation, autoimmune thyroiditis, appendicitis.

We evaluated incidence in the total population, i.e. did not require a “clean observation window” of individuals with data indicating they were free of any earlier manifestation of each condition during that window prior to the index start date, similar to the ACCESS protocol (5). This is also more consistent with real-life vaccine monitoring, as AESIs for most conditions may occur or be exacerbated in any individual, including those with earlier manifestations of the same pre-existing condition. The full ICD-10 definitions used are provided in Suppl Table 2.

### Analysis

The follow-up was evaluated for the full calendar year 2020, as well as for each quarter of 2020 (in Sweden, quarter 1 was essentially pre-pandemic, quarter 2 was first wave, quarter 3 was post-wave 1 and quarter 4 was second wave of the pandemic; the time course of the pandemic in cases and deaths is illustrated in Suppl Fig 1 as 7-day running average for COVID-19 cases and deaths over the year for 2020 from our database). For each event in each time period analysis (full year or quarter), individuals contributed person-time of follow-up from the start of that time period until the event occurred, end of the follow-up for that time period, or death, whichever occurred first.

We estimated incidence rates as the number of events divided by the person-time at risk (per 100,000 person-years). All estimates were weighted to the age and sex distribution of the total Swedish population using the sampling fractions for our population as weights. Using a similar approach as ref 7, we also calculated age/sex-specific incidence rates, where age was stratified in the age groups 1-5, 6-17, 18-35, 36-55, 56-64, 65-74, 75-84, and 85 years and older. All estimated incidence rates, overall and age/sex-specific rates, were classified using the World Health Organization Council for International Organizations of Medical Sciences (CIOMS) thresholds: very common (≥1/10), common (<1/10 to ≥1/100), uncommon (<1/100 to ≥1/1,000), rare (<1/1,000 to ≥1/10,000), and very rare (<1/10,000) (12).

Statistical analyses were performed using the R statistical program version 4.0.2 (13).

### Ethical approval

The research was approved by the Swedish Ethics Review Authority, No 2020-01800, 2020-05829, 2021-00267, 2021-00829, 2021-02106.

## Results

Overall, we had information on 972,723 study subjects (480,322 men and 492,401 women) who generated 957,477 person-years of follow-up during 2020 (Table 1). The corresponding weighted Swedish population sample is shown in Table 1. The full age and sex distribution of the study population, as well as sampling fractions from the Swedish total population, are shown in Suppl Table 1.

**Table 1.**
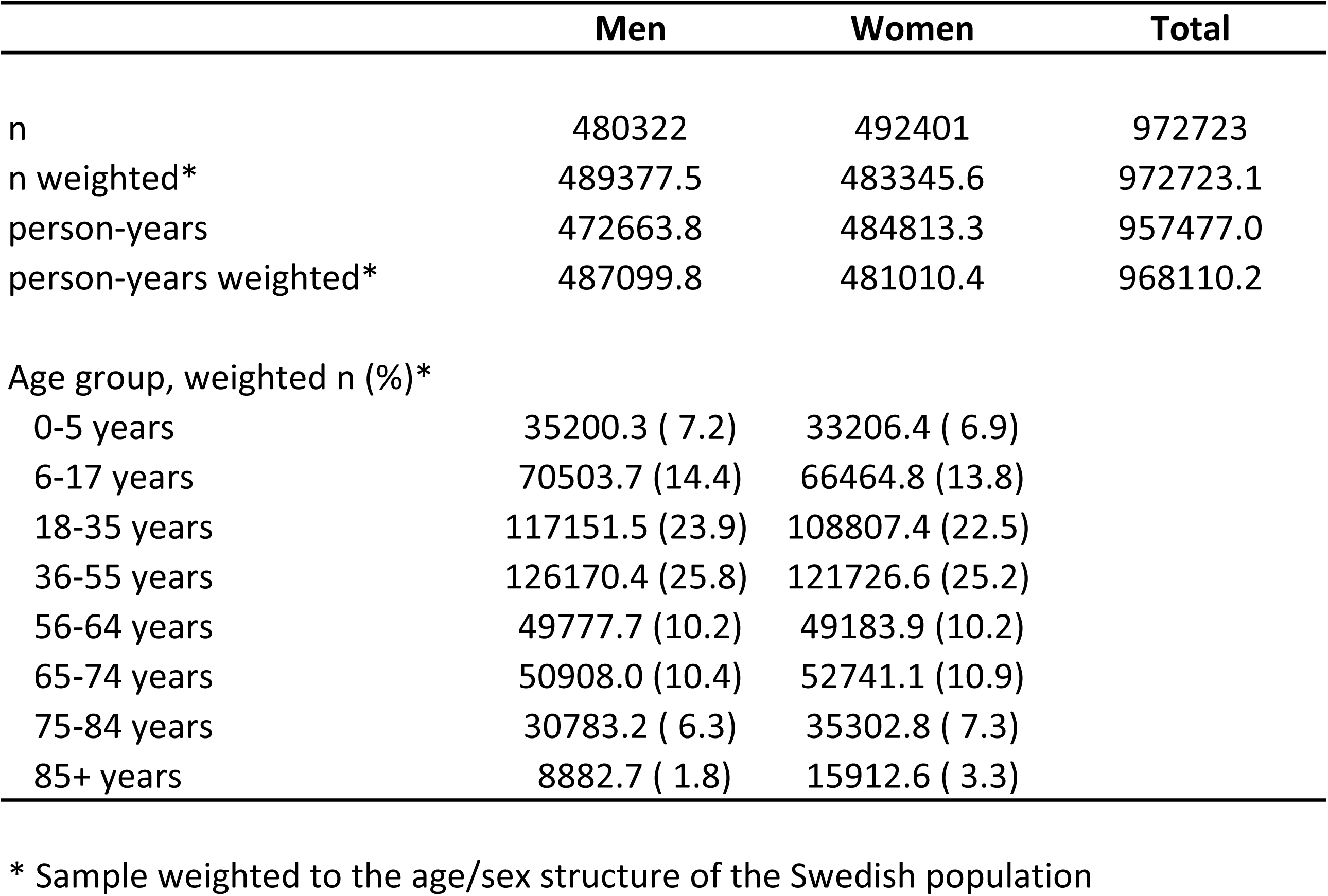
Age and sex distribution of the sampled Swedish population.

|  | Men | Women | Total |
| --- | --- | --- | --- |
| n | 480322 | 492401 | 972723 |
| n weighted* | 489377.5 | 483345.6 | 972723.1 |
| person-years | 472663.8 | 484813.3 | 957477.0 |
| person-years weighted* | 487099.8 | 481010.4 | 968110.2 |
| Age group, weighted n (%)* |  |  |  |
| 0-5 years | 35200.3 ( 7.2) | 33206.4 ( 6.9) |  |
| 6-17 years | 70503.7 (14.4) | 66464.8 (13.8) |  |
| 18-35 years | 117151.5 (23.9) | 108807.4 (22.5) |  |
| 36-55 years | 126170.4 (25.8) | 121726.6 (25.2) |  |
| 56-64 years | 49777.7 (10.2) | 49183.9 (10.2) |  |
| 65-74 years | 50908.0 (10.4) | 52741.1 (10.9) |  |
| 75-84 years | 30783.2 ( 6.3) | 35302.8 ( 7.3) |  |
| 85+ years | 8882.7 ( 1.8) | 15912.6 ( 3.3) |  |

\* Sample weighted to the age/sex structure of the Swedish population

The incidence rates of each AESI, overall, by sex, and stratified by age and sex are illustrated in Table 2. Each cell is colour-coded after the CIOMS adverse event frequency system (very common, common, uncommon, rare, or very rare).

**Table 2:** Age- and sex-stratified incidence rates per 100,00 person-years (with 95% confidence intervals), colour-coded by CIOMS frequency classification*.

|  |  | Age groups, years |  |  |  |  |  |  |  | total |
| --- | --- | --- | --- | --- | --- | --- | --- | --- | --- | --- |
|  | sex | 0-5 | 6-17 | 18-35 | 36-55 | 56-64 | 65-74 | 75-84 | 85+ |  |
| Arthritis (all types) | Female | 186.4 (137.2, 235.6) | 244.8 (205.4, 284.2) | 378.1 (337.4, 418.8) | 790.9 (735.8, 846.0) | 1428.2 (1317.2, 1539.1) | 1937.9 (1815.8, 2060.0) | 2204.0 (2071.0, 2337.1) | 1145.7 (1051.1, 1240.3) | 886.3 (859.3, 913.2) |
| Arthritis (all types) | Male | 247.1 (190.5, 303.7) | 159.8 (127.9, 191.8) | 151.7 (125.9, 177.5) | 385.6 (347.1, 424.0) | 739.4 (659.7, 819.2) | 998.2 (910.4, 1086.1) | 1279.2 (1176.1, 1382.2) | 804.1 (719.4, 888.8) | 450.6 (431.4, 469.8) |
| Myocardial infarction | Female | 0.000 (0.000, 0.000) | 0.000 (0.000, 0.000) | 2.23 (0.000, 5.34) | 27.3 (17.0, 37.6) | 125.7 (92.8, 158.6) | 336.3 (285.2, 387.4) | 704.2 (630.1, 778.3) | 1299.7 (1205.3, 1394.2) | 151.1 (141.7, 160.5) |
| Myocardial infarction | Male | 0.000 (0.000, 0.000) | 0.000 (0.000, 0.000) | 8.18 (2.11, 14.2) | 118.6 (97.2, 139.9) | 478.0 (414.1, 541.9) | 723.3 (648.6, 798.1) | 1161.6 (1064.0, 1259.2) | 1875.9 (1752.4, 1999.4) | 263.5 (250.2, 276.8) |
| Stroke | Female | 10.2 (0.000, 21.6) | 9.84 (1.96, 17.7) | 18.3 (9.31, 27.3) | 72.5 (55.7, 89.2) | 247.8 (201.5, 294.1) | 480.0 (419.0, 540.9) | 1143.0 (1049.2, 1236.9) | 2172.7 (2050.7, 2294.8) | 257.0 (244.7, 269.4) |
| Stroke | Male | 23.7 (6.14, 41.2) | 8.29 (1.02, 15.6) | 27.9 (16.9, 38.8) | 121.6 (99.9, 143.2) | 389.7 (331.9, 447.6) | 865.1 (783.2, 947.0) | 1776.5 (1658.2, 1894.7) | 2824.3 (2671.6, 2977.0) | 333.6 (319.0, 348.1) |
| Ischemic stroke | Female | 3.38 (0.000, 10.0) | 6.52 (0.128, 12.9) | 5.16 (0.624, 9.69) | 45.4 (32.1, 58.6) | 180.5 (141.0, 220.1) | 356.6 (304.0, 409.2) | 901.5 (817.8, 985.2) | 1772.0 (1661.5, 1882.5) | 195.2 (184.6, 205.8) |
| Ischemic stroke | Male | 6.76 (0.000, 16.1) | 3.25 (0.000, 7.76) | 18.4 (9.38, 27.5) | 86.3 (68.1, 104.5) | 310.8 (259.2, 362.4) | 654.5 (583.3, 725.8) | 1368.1 (1263.7, 1472.5) | 2097.7 (1965.6, 2229.8) | 251.5 (238.8, 264.1) |
| Acute kidney injury | Female | 3.38 (0.000, 10.0) | 1.61 (0.000, 4.75) | 17.9 (9.08, 26.7) | 40.4 (27.9, 52.9) | 92.3 (64.0, 120.5) | 259.5 (214.6, 304.4) | 707.0 (632.8, 781.2) | 1306.3 (1211.5, 1401.1) | 147.0 (137.7, 156.2) |
| Acute kidney injury | Male | 20.3 (4.06, 36.5) | 4.96 (0.000, 10.6) | 15.4 (7.29, 23.5) | 74.3 (57.4, 91.3) | 292.1 (241.9, 342.3) | 531.5 (467.3, 595.7) | 1163.6 (1067.8, 1259.4) | 2239.9 (2105.0, 2374.8) | 223.9 (212.1, 235.6) |
| Pulmonary embolism | Female | 0.000 (0.000, 0.000) | 9.63 (1.93, 17.3) | 46.6 (32.4, 60.8) | 72.0 (55.4, 88.7) | 178.1 (138.8, 217.3) | 341.9 (290.5, 393.4) | 619.7 (549.7, 689.8) | 657.2 (587.6, 726.7) | 152.3 (141.9, 162.7) |
| Pulmonary embolism | Male | 0.000 (0.000, 0.000) | 3.25 (0.000, 7.76) | 22.7 (12.7, 32.7) | 92.0 (73.2, 110.8) | 271.2 (222.9, 319.5) | 413.2 (356.6, 469.8) | 579.4 (510.3, 648.5) | 667.1 (592.2, 742.1) | 148.8 (138.3, 159.2) |
| Appendicitis | Female | 16.9 (2.09, 31.8) | 165.6 (133.2, 198.0) | 168.2 (141.4, 195.0) | 136.6 (113.7, 159.4) | 93.3 (64.7, 121.8) | 65.9 (43.4, 88.4) | 71.9 (47.1, 96.7) | 51.1 (31.0, 71.2) | 119.8 (109.4, 130.2) |
| Appendicitis | Male | 40.6 (17.6, 63.5) | 207.8 (171.2, 244.3) | 190.4 (161.9, 219.0) | 108.4 (88.1, 128.7) | 107.5 (77.1, 137.9) | 98.9 (71.2, 126.6) | 80.9 (54.5, 107.3) | 56.8 (34.2, 79.4) | 133.7 (122.5, 144.9) |
| ARDS | Female | 0.000 (0.000, 0.000) | 0.000 (0.000, 0.000) | 2.18 (0.000, 5.21) | 13.1 (5.96, 20.2) | 36.2 (18.4, 53.9) | 32.3 (16.4, 48.1) | 47.1 (27.6, 66.7) | 27.8 (15.0, 40.6) | 15.3 (11.9, 18.8) |
| ARDS | Male | 3.38 (0.000, 10.0) | 0.000 (0.000, 0.000) | 10.4 (3.57, 17.1) | 36.3 (24.4, 48.1) | 78.6 (52.6, 104.7) | 108.9 (79.9, 137.9) | 103.0 (75.0, 131.1) | 48.8 (28.6, 69.1) | 38.8 (33.1, 44.4) |
| Haemorrhagic stroke | Female | 6.77 (0.000, 16.2) | 1.71 (0.000, 5.06) | 13.2 (5.38, 21.0) | 30.1 (19.3, 40.9) | 74.1 (48.8, 99.4) | 126.9 (95.6, 158.2) | 263.3 (218.3, 308.2) | 413.4 (359.6, 467.2) | 65.5 (58.9, 72.1) |
| Haemorrhagic stroke | Male | 13.5 (0.271, 26.8) | 5.04 (0.000, 10.7) | 10.6 (4.03, 17.3) | 36.3 (24.4, 48.1) | 87.6 (60.1, 115.1) | 229.3 (187.1, 271.5) | 438.8 (380.3, 497.4) | 747.0 (668.0, 826.0) | 87.5 (80.0, 95.0) |
| Postinfectious arthritis | Female | 6.77 (0.000, 16.2) | 6.73 (0.133, 13.3) | 10.6 (4.00, 17.2) | 14.1 (6.70, 21.5) | 22.7 (8.61, 36.7) | 18.3 (6.33, 30.2) | 18.8 (6.97, 30.7) | 28.6 (14.1, 43.0) | 13.9 (10.5, 17.3) |
| Postinfectious arthritis | Male | 23.7 (6.14, 41.2) | 4.96 (0.000, 10.6) | 21.0 (11.5, 30.5) | 18.0 (9.67, 26.3) | 31.6 (15.0, 48.1) | 28.2 (13.4, 42.9) | 35.3 (18.0, 52.6) | 36.3 (19.0, 53.5) | 21.1 (16.8, 25.4) |
| Autoimmune thyroiditis | Female | 0.000 (0.000, 0.000) | 91.3 (67.4, 115.2) | 45.7 (31.3, 60.1) | 44.6 (31.6, 57.6) | 36.4 (18.6, 54.3) | 25.9 (11.8, 40.0) | 22.6 (8.46, 36.7) | 2.25 (0.000, 6.67) | 42.3 (36.1, 48.6) |
| Autoimmune thyroiditis | Male | 0.000 (0.000, 0.000) | 23.0 (11.0, 35.1) | 8.12 (2.09, 14.1) | 10.9 (4.45, 17.3) | 6.65 (0.000, 14.2) | 6.10 (0.000, 13.0) | 10.7 (1.08, 20.4) | 3.75 (0.000, 9.26) | 10.1 (7.06, 13.2) |
| Venous thromboembolism | Female | 0.000 (0.000, 0.000) | 3.21 (0.000, 7.66) | 11.8 (4.46, 19.1) | 22.9 (13.5, 32.3) | 47.6 (27.3, 68.0) | 44.5 (25.9, 63.1) | 88.4 (61.2, 115.5) | 54.7 (35.0, 74.3) | 26.8 (22.2, 31.4) |
| Venous thromboembolism | Male | 0.000 (0.000, 0.000) | 4.88 (0.000, 10.4) | 11.7 (4.43, 19.0) | 27.9 (17.6, 38.3) | 38.4 (20.1, 56.6) | 54.6 (34.0, 75.2) | 78.4 (53.0, 103.7) | 59.0 (36.5, 81.6) | 26.3 (21.6, 30.9) |
| Acute Liver failure | Female | 0.000 (0.000, 0.000) | 0.000 (0.000, 0.000) | 1.98 (0.000, 4.74) | 7.13 (1.85, 12.4) | 18.0 (5.52, 30.5) | 42.2 (24.1, 60.3) | 46.4 (27.1, 65.6) | 22.2 (9.03, 35.3) | 12.8 (9.66, 15.9) |
| Acute Liver failure | Male | 3.38 (0.000, 10.0) | 0.000 (0.000, 0.000) | 5.73 (0.698, 10.8) | 12.0 (5.20, 18.8) | 35.9 (18.3, 53.5) | 68.5 (45.5, 91.5) | 96.2 (67.6, 124.8) | 78.2 (51.7, 104.7) | 23.0 (18.8, 27.1) |
| Myocarditis | Female | 3.38 (0.000, 10.0) | 8.03 (0.992, 15.1) | 9.12 (2.77, 15.5) | 6.03 (1.20, 10.9) | 6.54 (0.000, 14.0) | 4.16 (0.000, 9.92) | 11.5 (2.14, 20.8) | 6.21 (1.24, 11.2) | 7.07 (4.59, 9.54) |
| Myocarditis | Male | 0.000 (0.000, 0.000) | 11.4 (2.95, 19.8) | 33.1 (21.4, 44.7) | 21.9 (12.7, 31.0) | 24.8 (10.1, 39.4) | 20.2 (7.68, 32.7) | 20.5 (7.41, 33.5) | 27.6 (12.3, 42.8) | 21.6 (17.2, 26.0) |
| Febrile seizure | Female | 261.1 (202.8, 319.3) | 3.42 (0.000, 8.15) | 0.000 (0.000, 0.000) | 0.000 (0.000, 0.000) | 0.000 (0.000, 0.000) | 0.000 (0.000, 0.000) | 1.66 (0.000, 4.92) | 0.000 (0.000, 0.000) | 18.5 (14.5, 22.6) |
| Febrile seizure | Male | 267.5 (208.6, 326.4) | 6.83 (0.137, 13.5) | 0.000 (0.000, 0.000) | 0.000 (0.000, 0.000) | 0.000 (0.000, 0.000) | 0.000 (0.000, 0.000) | 0.000 (0.000, 0.000) | 0.000 (0.000, 0.000) | 20.2 (15.9, 24.6) |
| Aseptic meningitis | Female | 0.000 (0.000, 0.000) | 1.61 (0.000, 4.75) | 11.0 (4.15, 17.9) | 0.971 (0.000, 2.88) | 0.000 (0.000, 0.000) | 0.000 (0.000, 0.000) | 0.000 (0.000, 0.000) | 0.000 (0.000, 0.000) | 2.94 (1.27, 4.61) |
| Aseptic meningitis | Male | 6.76 (0.000, 16.1) | 3.42 (0.000, 8.15) | 4.56 (0.081, 9.05) | 7.01 (1.82, 12.2) | 2.32 (0.000, 6.87) | 0.000 (0.000, 0.000) | 0.000 (0.000, 0.000) | 0.000 (0.000, 0.000) | 4.11 (2.09, 6.13) |
| Erythema multiforme | Female | 0.000 (0.000, 0.000) | 0.000 (0.000, 0.000) | 5.40 (0.643, 10.2) | 1.99 (0.000, 4.75) | 6.96 (0.000, 14.8) | 12.1 (2.42, 21.8) | 9.05 (1.01, 17.1) | 1.03 (0.000, 3.06) | 4.45 (2.51, 6.38) |
| Erythema multiforme | Male | 6.76 (0.000, 16.1) | 6.67 (0.132, 13.2) | 3.04 (0.000, 6.47) | 2.93 (0.000, 6.24) | 4.49 (0.000, 10.7) | 8.00 (0.161, 15.8) | 9.73 (0.932, 18.5) | 3.75 (0.000, 9.26) | 4.90 (2.87, 6.93) |
| Kawasaki's syndrome | Female | 20.3 (4.06, 36.6) | 4.92 (0.000, 10.5) | 0.000 (0.000, 0.000) | 0.000 (0.000, 0.000) | 0.000 (0.000, 0.000) | 0.000 (0.000, 0.000) | 0.000 (0.000, 0.000) | 0.000 (0.000, 0.000) | 2.07 (0.718, 3.43) |
| Kawasaki's syndrome | Male | 23.7 (6.14, 41.2) | 8.37 (1.03, 15.7) | 2.02 (0.000, 4.83) | 0.000 (0.000, 0.000) | 0.000 (0.000, 0.000) | 0.000 (0.000, 0.000) | 0.000 (0.000, 0.000) | 0.000 (0.000, 0.000) | 3.39 (1.62, 5.17) |
| MISC | Female | 0.000 (0.000, 0.000) | 6.63 (0.130, 13.1) | 0.000 (0.000, 0.000) | 0.000 (0.000, 0.000) | 0.000 (0.000, 0.000) | 0.000 (0.000, 0.000) | 1.66 (0.000, 4.92) | 0.000 (0.000, 0.000) | 1.03 (0.108, 1.96) |
| MISC | Male | 3.38 (0.000, 10.0) | 0.000 (0.000, 0.000) | 0.000 (0.000, 0.000) | 0.000 (0.000, 0.000) | 2.32 (0.000, 6.87) | 2.00 (0.000, 5.92) | 2.55 (0.000, 7.55) | 0.000 (0.000, 0.000) | 0.848 (0.007, 1.69) |
| Disseminated intravascular coagulation | Female | 0.000 (0.000, 0.000) | 3.31 (0.000, 7.91) | 1.19 (0.000, 3.51) | 0.000 (0.000, 0.000) | 0.000 (0.000, 0.000) | 3.93 (0.000, 9.39) | 0.000 (0.000, 0.000) | 1.03 (0.000, 3.06) | 1.19 (0.170, 2.20) |
| Disseminated intravascular coagulation | Male | 0.000 (0.000, 0.000) | 1.71 (0.000, 5.05) | 1.22 (0.000, 3.63) | 0.000 (0.000, 0.000) | 2.17 (0.000, 6.41) | 2.05 (0.000, 6.06) | 1.54 (0.000, 4.57) | 0.000 (0.000, 0.000) | 1.07 (0.090, 2.05) |

\* CIOMS: Council of International Organizations of Medical Sciences

| CIOMS Frequency classification |
| --- |
| Very rare: <1/10,000 |
| Rare: >1/10,000 AND <1/1,000 |
| Uncommon: >1/1,000 AND <1/100 |
| Common: >1/100 AND <1/10 |
| Very common: >1/10 |

There is substantial variation for most AESIs by age, while sex differences are more discrete for most AESIs. The age variation is clearly seen in Figure 1, where several common conditions increase as expected with age, but some conditions show a different pattern. Appendicitis, aseptic meningitis, autoimmune thyroiditis, Kawasaki syndrome and MISC are more common in children or younger ages. Myocarditis, DIC and erythema multiforme have a more equal distribution across ages. The greatest sex differences were seen for myocarditis, arthritis, autoimmune thyroiditis and ARDS.

**Figure 1:**
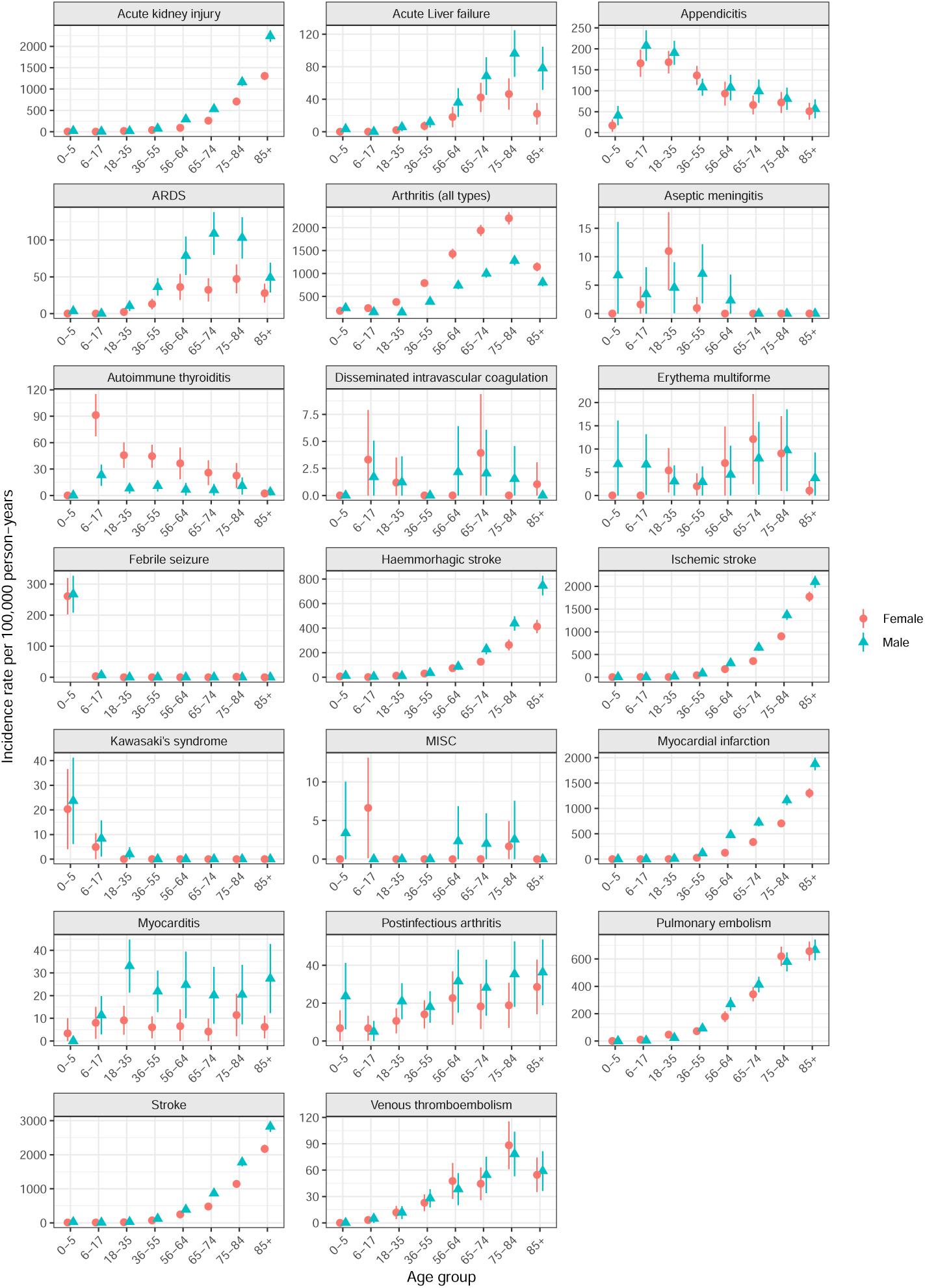
Age- and sex-stratified incidence rates of 14 adverse events of special interest (AESIs) per 100,000 person-years (with 95% confidence intervals) in the Swedish population

In Figure 2, the incidence trend over the four quarters of 2020 is illustrated. Some AESIs, e.g. venous thromboembolism, hemorrhagic stroke and postinfectious arthritis exhibit a decline in incidence during the year. Some AESIs, especially ARDS and acute liver failure follow a trend with highest incidence in the second and fourth quarter, corresponding approximately to wave 1 and wave 2 of the COVID-19 pandemic in Sweden. Erythema multiforme shows an inverse pattern, being lower during the wave 1 and 2 quarters. The MISC diagnosis was not apparent or little used before quarter 4, where the related diagnosis Kawasaki syndrome tended to decrease after an earlier increase during the first pandemic quarter.

**Figure 2:**
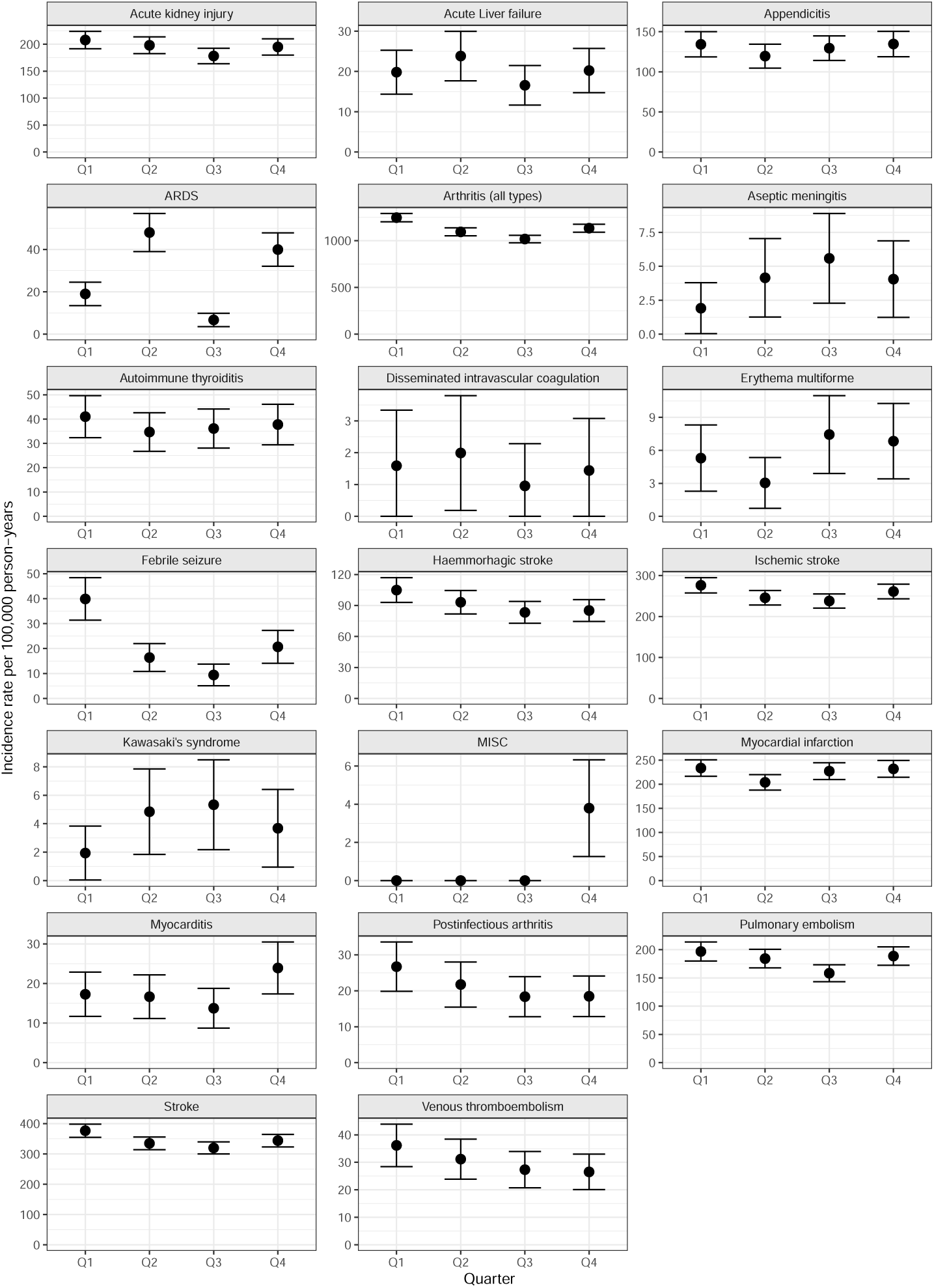
Incidence rates of 14 adverse events of special interest (AESIs) per 100,000 person-years (with 95% confidence intervals) in the Swedish population, by quarter of the year 2020.

## Discussion

We report descriptive epidemiology for a number of AESIs of potential interest for safety follow-up of COVID-19 vaccines, from an unselected national population. We describe baseline rates of these AESIs during the year 2020 overall and by quarter. Our results illustrate a number of important points for upcoming studies of vaccine safety.

First, the substantial variability of rates by age, and for certain outcomes also by sex, imply that it will be important to have adequate demographic comparability for vaccinated and non-vaccinated groups that are compared, or alternatively to ensure adequate adjustment or standardization for age and sex if background rates are to be used for safety surveillance. It also means that adverse event rates reported without reference to age or sex distributions will be very difficult to interpret.

Second, the clear trends over quarters seen for some AESI during the pandemic in Sweden, with variable patterns, imply that comparing rates across appropriate time periods will be crucial. For example, a doubled risk of an adverse event by a vaccine during a period when in fact the background rate of that event in the underlying population as measured by healthcare encounters is temporarily half of what it normally is may not be detected if a comparison is made with a historical or other period when the rate is at its habitual level. Similarly, for events that are increased by the infectious disease itself, e.g. MISC, if these are also increased by a vaccine, this may be difficult to discern unless the comparison is made during contemporaneous time periods when the disease is active. Therefore, contemporaneous studies such as the current study are essential, and it will not be adequate to rely on historical background rates from literature only. Further, we found substantial heterogeneity across time periods *during* the early phases of the pandemic that vaccines are now used to combat, suggesting that using a single overall estimate from one selected time period may be inadequate to represent the true comparator event incidence. Such patterns may potentially be related to COVID-19 pandemic waves (i.e. background incidence of COVID-19), seasonal variations, healthcare system overload or other healthcare-related changes during a time of pandemic, e.g. changes in registration or reporting of diagnoses related to the pandemic, changing consultation behaviour of patients, increasing digital healthcare delivery, or other factors.

For example, coagulation disorders have been associated with COVID-19 (14), and potential adverse events related to coagulation disorders have recently been linked to some COVID-19 vaccines (15-18). Here we report background rates for venous thromboembolism, pulmonary embolism, stroke and ischemic stroke and DIC, conditions that are all relevant for this AESI category. It may be noted that rates for these individual conditions have different relationships with age, as well as across the 4 quarters that we studied. Myocarditis is another condition that has recently been of interest as a potential vaccine adverse event, particularly in younger males (19), and has been related to COVID-19 (20). In our data, we see substantially higher background rates in men than women, and with a tendency that the highest rates are observed in the 18-35 year age group, which will be important to consider when further evaluating this potential safety issue. The rate of myocarditis was also highest in the 4^th^ quarter, during the intensive 2^nd^ pandemic wave in Sweden, so background incidence of COVID-19 over time may be quite important to consider.

Overall, it is comforting that despite variability and differences, our findings also show consistency with prior reports from the literature on many points. Nevertheless, recent large multi-country studies have shown considerable heterogeneity across populations, for example data reported from the ACCESS project, which showed similar magnitudes of heterogeneity in background rates (3). The pooled rates across databases from a study by the OHDSI group showed many similarities to our rates for AESIs that were included in both studies; in addition, they also reported variability across sites and databases (8). This further underscores that population heterogeneity needs to be taken into account in the evaluation of AESIs.

Considering the large variability that was seen across age, sex, and time period, we would urge caution when using external reference incidence rates to make comparisons with incidences observed in vaccinated individuals, as the risk of systematic bias due to differences between populations and situations may be substantial. Comparing incidence rates that originate from different data sources may carry additional risk of systematic bias due to data-related differences between data sources. Indeed, large variations have been seen between different types of data such as electronic health records and claims data sources even when using harmonized type of analysis and outcome definitions (8). Using rates from randomised trials or spontaneous reporting data may induce even greater variability, related to the specific data collection mechanisms for these activities. Overall, external rates from various sources will continue to be one important source of comparator and contextual information for spontaneous adverse event reporting and estimates of outcome rates in vaccinated groups of people (21-23). But this should not be considered sufficient. Formal studies with contemporaneous comparators, in different databases and using appropriate carefully tuned epidemiological study designs, will be essential components of adverse event monitoring efforts. Fortunately, we are increasingly in a position to do this, as illustrated by the SCIFI-PEARL study initiative (9) and other large-scale register data initiatives.

A limitation of this observational study in common with most, is that all outcomes may be subject to measurement error. As all outcome definitions are based on the presence of specific ICD-10 codes and were not further validated, they may lack in sensitivity or specificity, which may affect the estimated incidence rates. However, as a reflection of actual healthcare encounters in the Swedish healthcare system, many of these codes and algorithms have been shown to have good validity (10, 24). Interestingly, the existence of different patterns of misclassification in different datasets is in fact a strong argument to use the same data source, population and time period for both groups compared when conducting AESI evaluation. The analysis relied on data from 2020 using a target population that was a defined random sample of the total Swedish population on 1 Jan 2020. This provides our analysis with a good level of generalizability. We did not exclude individuals with previous events of the same kind as the AESIs. This design choice is aligned with real-life concerns where AESIs will be an issue whether they occur in people with or without underlying or pre-existing conditions. For many conditions, rates of events are generally somewhat higher in individuals with underlying comorbidity of the same type, but in most cases this difference is not very large and the group with underlying comorbidity is a small minority of the total population, so that rates estimated after excluding them will often only be marginally lower than rates estimated from the total population. When using background incidence rates for comparison with rates in vaccinated individuals it is also important to recognize that some AESIs may overlap with symptoms of the infection targeted by the vaccines (in this case COVID-19), which will complicate the comparisons, since the rate of infection and/or infection-related symptoms will be lower in the vaccinated individuals if the vaccine is efficacious. Finally, in this study we have estimated incidence rates in sub-categories of age, sex and time periods, and random variation due to small numbers should be considered when interpreting the results. Nevertheless, our study overall is large and we present rates “as observed” i.e. descriptively, which incorporates actual random variation in the underlying data and diseases.

In summary, this study, which is based on high-quality Swedish healthcare register data, demonstrates that there is a large variation in estimated AESI incidence rates by age and sex, as well as time period in the Swedish population during 2020. While baseline rates continue to be a vital reference point and analytic tool for Covid-19 vaccine monitoring (22), these results emphasize the need for well-designed analytical studies with attention to stratification, standardization or adjustment for age, sex and time period. These background rates provide useful real-world clinical context for activities aiming at vaccine monitoring and ensuring patient safety as Covid-19 vaccines are applied to combat the pandemic worldwide.

## Data Availability

The data used for this article are subject to Swedish confidentiality and personal data legislation and restricted in use to the scope of ethical approvals. Researchers interested in collaborating around analyses are requested to contact the authors.

## Supplementary Materials

Supplementary Table 1: Study population by age and sex, numbers and sampling fractions from Swedish total population

Supplementary Table 2. Definitions of adverse events of interest (AESIs)

## Acknowledgements

We acknowledge Jonatan Nåtman for statistical analysis support.

## References

1. Huang C, Wang Y, Li X, et al. Clinical features of patients infected with 2019 novel coronavirus in Wuhan, China. The Lancet. 2020;395 (10223):497–506. doi:10.1016/S0140-6736(20)30183-5.

2. World Health Organization. Coronavirus disease (Covid-19) Weekly Epidemiological Update 5 January 2020. Available from: https://www.who.int/emergencies/diseases/novel-coronavirus-2019/situation-reports/. accessed 6 January 2021.

3. Ludvigsson JF. The first eight months of Sweden’s COVID-19 strategy and the key actions and actors that were involved. Acta Paediatr. 2020 Dec;109(12):2459–2471. doi: 10.1111/apa.15582. Epub 2020 Oct 11..

4. Tegnell A. The Swedish public health response to COVID-19. APMIS. 2021 Jul;129(7):320–323. doi: 10.1111/apm.13112.

5. ACCESS: Background rates of Adverse Events of Special Interest for monitoring COVID-19 vaccines. 2020. http://www.encepp.eu/encepp/viewResource.htm?id=37274 (accessed 13 Sept 2021).

6. Center for Biologics Evaluation and Research Office of Biostatistics and Epidemiology. CBER Surveillance Program Background Rates of Adverse Events of Special Interest for COVID-19 Vaccine Safety Monitoring Protocol. https://www.bestinitiative.org/wp-content/uploads/2021/02/C19-Vaccine-Safety-AESI-Background-Rate-Protocol-FINAL-2020.pdf (accessed 13 Sept 2021).

7. Report of CIOMS Working Group on Vaccine Safety. CIOMS Guide to Active Vaccine Safety Surveillance. https://cioms.ch/wp-content/uploads/2020/04/240WEB-CIOMS-Guide-AVSS-20170202-protected.pdf (accessed March 11, 2021).

8. Li X, Ostropolets A, Makadia R, Shoaibi A, Rao G, Sena AG, Martinez-Hernandez E, Delmestri A, Verhamme K, Rijnbeek PR, Duarte-Salles T, Suchard M, Ryan P, Hripcsak G, Prieto-Alhambra D. Characterising the background incidence rates of adverse events of special interest for covid-19 vaccines in eight countries: multinational network cohort study BMJ 2021; 373 :n1435 doi:10.1136/bmj.n1435.

9. Nyberg F, Franzén S, Lindh M, Vanfleteren L, Hammar N, Wettermark B, Sundström J, Santosa A, Björck S, Gisslén M. Swedish Covid-19 Investigation for Future Insights - A Population Epidemiology Approach Using Register Linkage (SCIFI-PEARL). Clin Epidemiol. 2021;13:649–659. doi: 10.2147/CLEP.S312742.

10. Ludvigsson JF, Andersson E, Ekbom A, Feychting M, Kim JL, Reuterwall C, Heurgren M, Olausson PO. External review and validation of the Swedish national inpatient register. BMC Public Health. 2011 Jun 9;11:450. doi: 10.1186/1471-2458-11-450.

11. Brighton Collaboration. Priority list of adverse events of special interest: COVID-19. 2021. https://brightoncollaboration.us/covid-19/; https://brightoncollaboration.us/wp-content/uploads/2021/01/COVID-19-updated-AESI-list.pdf (accessed 14 Sept, 2021).

12. The Council for International Organizations of Medical Sciences (CIOMS). Guidelines for Preparing Core Clinical-Safety Information on Drugs Second Edition – Report of CIOMS Working Groups III and V. Geneva, 1999.

13. R Core Team (2020). R: A language and environment for statistical computing. R Foundation for Statistical Computing, Vienna, Austria. https://www.R-project.org/.

14. Leentjens J, van Haaps TF, Wessels PF, Schutgens REG, Middeldorp S. COVID-19-associated coagulopathy and antithrombotic agents-lessons after 1 year. Lancet Haematol. 2021 Jul;8(7):e524–e533. doi: 10.1016/S2352-3026(21)00105-8. Epub 2021 Apr 27.

15. Wise J. Covid-19: European countries suspend use of Oxford-AstraZeneca vaccine after reports of blood clots. BMJ 2021; 372: n699.

16. Østergaard SD, Schmidt M, Horváth-Puhó E, Thomsen RW, Sørensen HT. Thromboembolism and the Oxford-AstraZeneca COVID-19 vaccine: side-effect or coincidence? Lancet. 2021 Apr 17;397(10283):1441–1443. doi: 10.1016/S0140-6736(21)00762-5. Epub 2021 Mar 30.

17. Marcucci R, Marietta M. Vaccine-induced thrombotic thrombocytopenia: the elusive link between thrombosis and adenovirus-based SARS-CoV-2 vaccines. Intern Emerg Med. 2021 Aug;16(5):1113–1119. doi: 10.1007/s11739-021-02793-x. Epub 2021 Jun 30.

18. European Medicines Agency. EMA raises awareness of clinical care recommendations to manage suspected thrombosis with thrombocytopenia syndrome. News 07/06/2021. https://www.ema.europa.eu/en/news/ema-raises-awareness-clinical-care-recommendations-manage-suspected-thrombosis-thrombocytopenia. (accessed 14 Sept, 2021).

19. European Medicines Agency. Comirnaty and Spikevax: possible link to very rare cases of myocarditis and pericarditis. News 09/07/2021. https://www.ema.europa.eu/en/news/comirnaty-spikevax-possible-link-very-rare-cases-myocarditis-pericarditis. (accessed 14 Sept, 2021).

20. Boehmer TK, Kompaniyets L, Lavery AM, Hsu J, Ko JY, Yusuf H, Romano SD, Gundlapalli AV, Oster ME, Harris AM. Association Between COVID-19 and Myocarditis Using Hospital-Based Administrative Data - United States, March 2020-January 2021. MMWR Morb Mortal Wkly Rep. 2021 Sep 3;70(35):1228–1232. doi: 10.15585/mmwr.mm7035e5.

21. Black S, Eskola J, Siegrist C-A, et al. Importance of background rates of disease in assessment of vaccine safety during mass immunisation with pandemic H1N1 influenza vaccines. Lancet 2009; 374: 2115–22.

22. Rasmussen TA, Jørgensen MRS, Bjerrum S, et al. Use of population based background rates of disease to assess vaccine safety in childhood and mass immunisation in Denmark: nationwide population based cohort study. BMJ 2012; 345: e5823.

23. Black SB, Law B, Chen RT, et al. The critical role background rates of possible adverse events in the assessment of COVID-19 vaccine safety. Vaccine 2021; published online March. DOI:10.1016/j.vaccine.2021.03.016.

24. Friberg L, Skeppholm M. Usefulness of Health Registers for detection of bleeding events in outcome studies. Thromb Haemost 2016; 116(06): 1131–1139. DOI: 10.1160/TH16-05-0400.

