## Supplemental tables and figures for "Adverse events of special interest for COVID-19 vaccines - background incidences vary by sex, age and time period and are affected by the pandemic"

**Supplementary Table 1: Study population sample by age and sex, numbers and sampling fractions from Swedish total population**

| Age group, years | Men, sampled n | Men, sampled proportion | Women, sampled n | Women, sampled proportion | Total, sampled n | Total, sampled proportion |
|---|---|---|---|---|---|---|
| 0-4 | 25000 | 0.081 | 25000 | 0.086 | 50000 | 0.083 |
| 5-9 | 25000 | 0.078 | 25000 | 0.082 | 50000 | 0.080 |
| 10-14 | 25000 | 0.080 | 25000 | 0.085 | 50000 | 0.082 |
| 15-19 | 25000 | 0.084 | 25000 | 0.092 | 50000 | 0.088 |
| 20-24 | 25000 | 0.080 | 25000 | 0.090 | 50000 | 0.085 |
| 25-29 | 25000 | 0.066 | 25000 | 0.070 | 50000 | 0.068 |
| 30-34 | 25000 | 0.068 | 25000 | 0.072 | 50000 | 0.070 |
| 35-39 | 25000 | 0.075 | 25000 | 0.079 | 50000 | 0.077 |
| 40-44 | 25000 | 0.078 | 25000 | 0.081 | 50000 | 0.079 |
| 45-49 | 25000 | 0.074 | 25000 | 0.076 | 50000 | 0.075 |
| 50-54 | 25000 | 0.073 | 25000 | 0.075 | 50000 | 0.074 |
| 55-59 | 25000 | 0.080 | 25000 | 0.081 | 50000 | 0.081 |
| 60-64 | 25000 | 0.088 | 25000 | 0.088 | 50000 | 0.088 |
| 65-69 | 25000 | 0.094 | 25000 | 0.092 | 50000 | 0.093 |
| 70-74 | 25000 | 0.091 | 25000 | 0.087 | 50000 | 0.089 |
| 75-79 | 25000 | 0.122 | 25000 | 0.112 | 50000 | 0.117 |
| 80-84 | 25000 | 0.206 | 25000 | 0.165 | 50000 | 0.183 |
| 85-89 | 25000 | 0.391 | 25000 | 0.251 | 50000 | 0.306 |
| 90-94 | 24740 | 1 | 25000 | 0.482 | 49740 | 0.649 |
| 95+ | 5580 | 1 | 17403 | 1 | 22983 | 1 |
|  | 480320 |  | 492403 |  | 972723 |  |

**Supplementary Table 2. Definitions of adverse events of interest (AESIs)**

| **Outcome** | **ICD-10 codes** |
|---|---|
| Aseptic meningitis | A87 |
| | B261 |
| | G030 |
| Febrile seizure | R560 |
| Kawasaki's syndrome | M303 |
| Multisystem inflammatory syndrome in children (MISC) | U109 |
| Postinfectious arthritis | M015 |
| | M018 |
| | M02 |
| | M03 |
| Arthritis, broad (all types) | M015 |
| | M018 |
| | M02 |
| | M03 |
| | M05 |
| | M06 |
| | M07 |
| | M08 |
| | M09 |
| | M13 |
| Myocarditis | I012 |
| | I090 |
| | I40 |
| | I41 |
| | I514 |
| Acute respiratory distress syndrome (ARDS) | J80 |
| | P220 |
| Myocardial infarction, broad | I21 |
| | I22 |
| | I23 |

| | |
|---|---|
| Stroke | F010 |
| | F011 |
| | F013 |
| | I63 |
| | P101 |
| | P524 |
| | P526 |
| | I60 |
| | I61 |
| | I62 |
| | I64 |
| Ischemic stroke | F010 |
| | F011 |
| | F013 |
| | I63 |
| Haemmorhagic stroke | P101 |
| | P524 |
| | P526 |
| | I60 |
| | I61 |
| | I62 |
| Venous thromboembolism | I822 |
| | I823 |
| | I828 |
| | I829 |
| Pulmonary embolism | I26 |
| Acute kidney injury | N17 |
| | O904 |
| Acute Liver failure | K720 |
| | K729 |
| Erythema multiforme | L51 |
| Disseminated intravascular coagulation | D65 |
| | P60 |
| Autoimmune thyroiditis | E063 |
| Appendicitis | K35 |
| | K36 |
| | K37 |

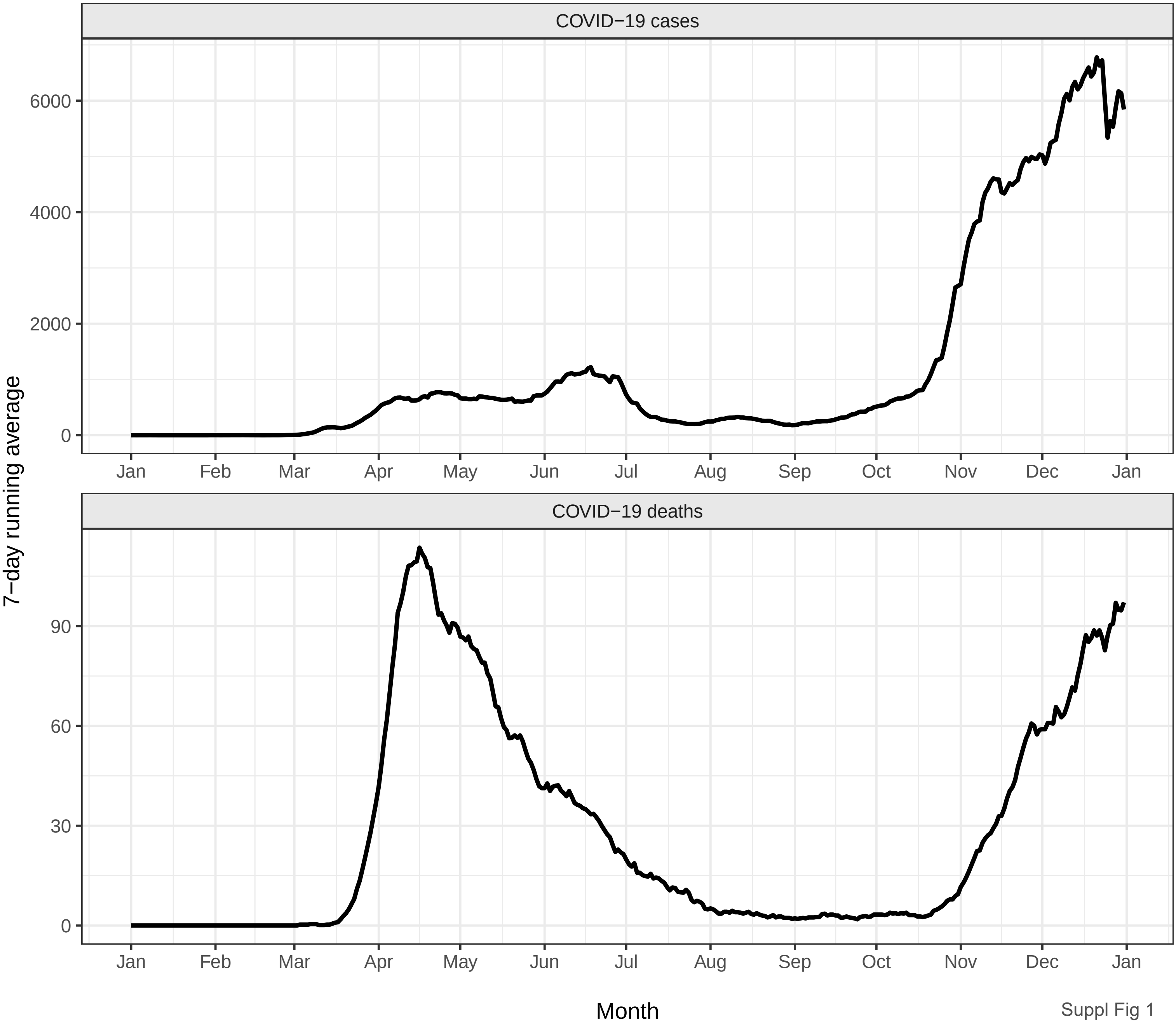
COVID−19 cases
6000
4000
2000
0
Jan
Feb
Mar
Apr
May
Jun
Jul
Aug
Sep
Oct
Nov
Dec
Jan
COVID−19 deaths
90
60
30
0
7−day running average
Month

Suppl Fig 1
